# Alleviation of allergic rhinoconjunctivitis symptoms in participants treated with a 0.005% tacrolimus eye drop solution

**DOI:** 10.1101/2024.04.30.24306626

**Authors:** S. Sladek, N. Unger-Manhart, C. Siegl, H. Dellago, P. Zieglmayer, P. Lemell, M. Savli, R. Zieglmayer, W. Geitzenauer, M. Laengauer, E. Prieschl-Grassauer

## Abstract

**Purpose:** This randomized, placebo-controlled, crossover, double-blind, single site trial was aimed to evaluate efficacy and safety of Tacrosolv, a novel eye drop solution containing solubilized tacrolimus, in adult participants with grass pollen induced allergic conjunctivitis.

**Methods:** 64 adult participants with proven grass pollen allergy were randomized to either 2.5 µg or 5 µg tacrolimus/eye/day or placebo treatment for 8 days. Allergic symptoms were induced by 4h grass pollen exposure on day 1 and day 8. After a 2-week wash-out period, placebo participants crossed over to high or low dose and vice versa, and repeated treatment and exposure. During exposure, participants recorded ocular, nasal and respiratory allergy symptoms every 15 minutes. The primary endpoint was the mean ‘Total Ocular Symptom Score’ (TOSS) on Day 8. Objective ocular safety parameters were assessed before, during and after exposure. Adverse events (AEs) were recorded throughout the study.

**Results:** On Day 8, TOSS was reduced towards the end of exposure in participants receiving Tacrosolv high dose compared to placebo (p<0.05 at timepoints 3h45min and 4h). Accordingly, intensity of the single ocular symptoms like redness and watery eyes was reduced versus placebo by end of exposure on Day 8. A 26% reduction of baseline adjusted TOSS from day 1 to day 8 was observed in participants treated with high dose Tacrosolv, whereas placebo treated participants showed no difference in TOSS between day 1 and day 8. Interestingly, a significant reduction of total nasal symptoms, mainly itching and sneezing, was seen both on day 1 and day 8 in participants treated with high dose Tacrosolv (p<0.05). No safety concerns were raised upon ocular assessments by the investigator like redness of the eye, corneal and conjunctival staining. All AEs were resolved within the study period.

**Conclusion:** Treatment with Tacrosolv at the dose and frequency studied is safe and alleviates symptoms in participants suffering from allergic rhinoconjunctivitis.

**Trial registration:** NCT04532710; EudraCT No. 2019-002847-62

## Introduction

Tacrolimus is a macrolide lactam that acts as immunosuppressant by inhibiting T-lymphocyte signal transduction and mast cell function^1,2^ by suppressing cytokine and histamine release and impairing prostaglandin synthesis.^3,4^ Tacrolimus is widely used after organ transplantation to reduce the risk of organ rejection, as well as in chronic inflammatory conditions of the skin.^5^ More recently, clinical trials have been undertaken applying tacrolimus in ocular diseases like corneal graft rejection, herpetic stromal keratitis, inflammatory conjunctival and corneal diseases, and uveitis.^6,7^ There is a long-term established off-label use of systemically applied tacrolimus for the treatment of non-infectious uveitis posterior, as well as ocular application of tacrolimus (mainly off-label use of the intravenous tacrolimus product Prograf^®^ and the tacrolimus ointment Protopic^®^) in patients with (refractory) vernal keratoconjunctivitis (VKC), (intractable) allergic (kerato)conjunctivitis, (refractory) atopic keratoconjunctivitis (AKC), contact lens-induced papillary conjunctivitis and graft-versus-host-disease (GVHD) as well as allergic conjunctival granuloma and Splendore-Hoeppli phenomenon documented. Tacrolimus is currently available only as suspension or emulsion, since the substance’s highly hydrophobic character (water solubility: 5–8 μg/mL) and high molecular weight (804.02 g/mol) have so far precluded development of a tacrolimus solution. The currently available dosage forms do not allow the substance to effectively penetrate the cornea and conjunctiva and reach effective therapeutic intraocular concentrations. This has so far hindered delivery of its full therapeutic potential for use in immunologic eye diseases. Currently, there is only one ophthalmic formulation of tacrolimus, marketed as Talymus^®^ in Japan and South Korea for the treatment of vernal keratoconjunctivitis (Senju Pharmaceutical Co., Ltd., Osaka, Japan). Talymus^®^ is a suspension with a tacrolimus concentration of 0.1% (1 mg/ml).

Allergic conjunctivitis is one of the most common comorbidities of allergic diseases, especially of allergic rhinitis. Rhinoconjunctivitis is an allergic condition of the nasal mucosa and the eyes. Allergic conjunctivitis is triggered by hypersensitivity to certain pollens and other airborne allergens and causes several symptoms such as red eyes, itchy eyes, watery eyes and a scratchy feeling in the eye. It is clinically defined as a symptomatic disorder induced by immunoglobulin E (IgE)-mediated inflammation after allergen exposure to the conjunctiva of the eye. Allergen-bound IgE on the surface of mast cells induces mast cell degranulation and release of allergic and inflammatory mediators such as histamines, leukotrienes, prostaglandin D2, tryptase, kinins, and pro-inflammatory cytokines such as TNF alpha.^8^

Environmental exposure chambers allow exposing allergic study participants to a physiological allergen challenge comparable to real-life allergen exposure, in contrast to intranasal and intraocular challenge by direct application of challenge solutions. The Vienna Challenge Chamber (VCC) has offered the opportunity to obtain assessments of allergic responses in allergic subjects in a several hours period, with high reproducibility, resulting in a rather small number of patients necessary to obtain significant results.

Tacrolimus eye drops (Tacrosolv) contain 50 µg/mL (0.005%) tacrolimus dissolved in our proprietary Marinosolv^®^ formulation. We have previously demonstrated the bioavailability and permeability of solubilized tacrolimus when applying Tacrosolv topically in an *ex vivo* as well as *in vivo* animal model.^9^ For the intended clinical purpose, a concentration of 50 µg/mL (0.005%; with a maximum recommended dose of maximal two drops per eye (maximal daily dose of 5 µg tacrolimus per eye) was chosen based on *ex vivo* experiments with porcine eyes, where a similar volume (50 µL) of Tacrosolv (5 µg tacrolimus /eye) and of Talymus^®^ (50 µg tacrolimus /eye) resulted in similar conjunctival drug concentrations (data not shown).

The goal of the study presented here was to establish proof of concept for the efficacy and safety of Tacrosolv for the treatment of inflammatory-driven ophthalmic diseases, using allergen exposure challenge as a simple, quick and controllable model system. This is the first clinical study using the proprietary Tacrosolv formulation.

## Methods

### Study Design

This was a randomized, placebo-controlled, crossover, double-blind, single site trial in adult participants (18-65 years of age) with documented grass specific Immunglobulin E (IgE) reactivity and a history of grass pollen induced rhinoconjunctivitis with or without controlled asthma. Two dose groups, namely low dose (2.5 µg tacrolimus/eye/day) and high dose (5 µg tacrolimus/eye/day) were evaluated during two treatment periods with a duration of 8 days each. The crossover design ensured that individual participants received either the low dose or the high dose of Tacrosolv in one treatment period and placebo in the other treatment period.

At screening (visit 1), medical and allergic history, safety lab as well as inclusion and exclusion criteria were retrieved, and all safety assessments were conducted. At least one week prior to the first treatment period (visit 2), participants were screened for appropriate allergic response during a grass pollen challenge chamber session.

At visit 3 (Day 1 of treatment period [TMP] 1), eligible participants were randomly assigned to one of the four treatment arms in a fully blinded fashion. See graphical abstract in **Figure 1** for treatment arms. After positive completion of all study relevant assessments, baseline values for symptom scores were assessed and participants were administered their first treatment 30 minutes before entering the challenge chamber. The total ocular symptom score (TOSS), total nasal symptom score (TNSS) and total respiratory symptom score (TRSS) as well as nasal airflow (AAR) and lung function were assessed at defined timepoints during exposure. Objective ocular assessments were performed before and after the provocation session. After the allergen exposure, participants received study medication for the home treatment phase (days 2 to 7) and continued administration until day 7.

**Figure 1:**
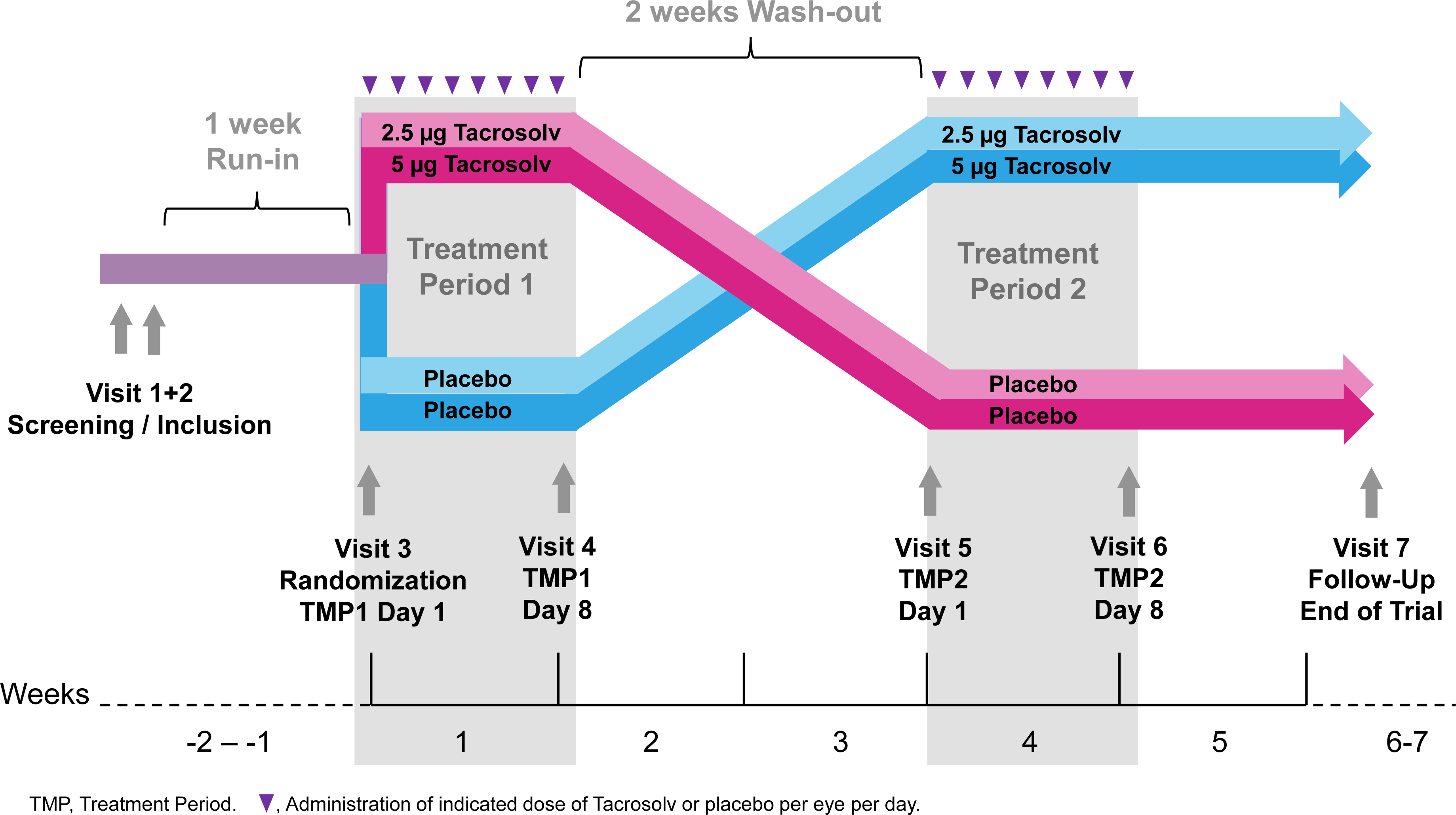
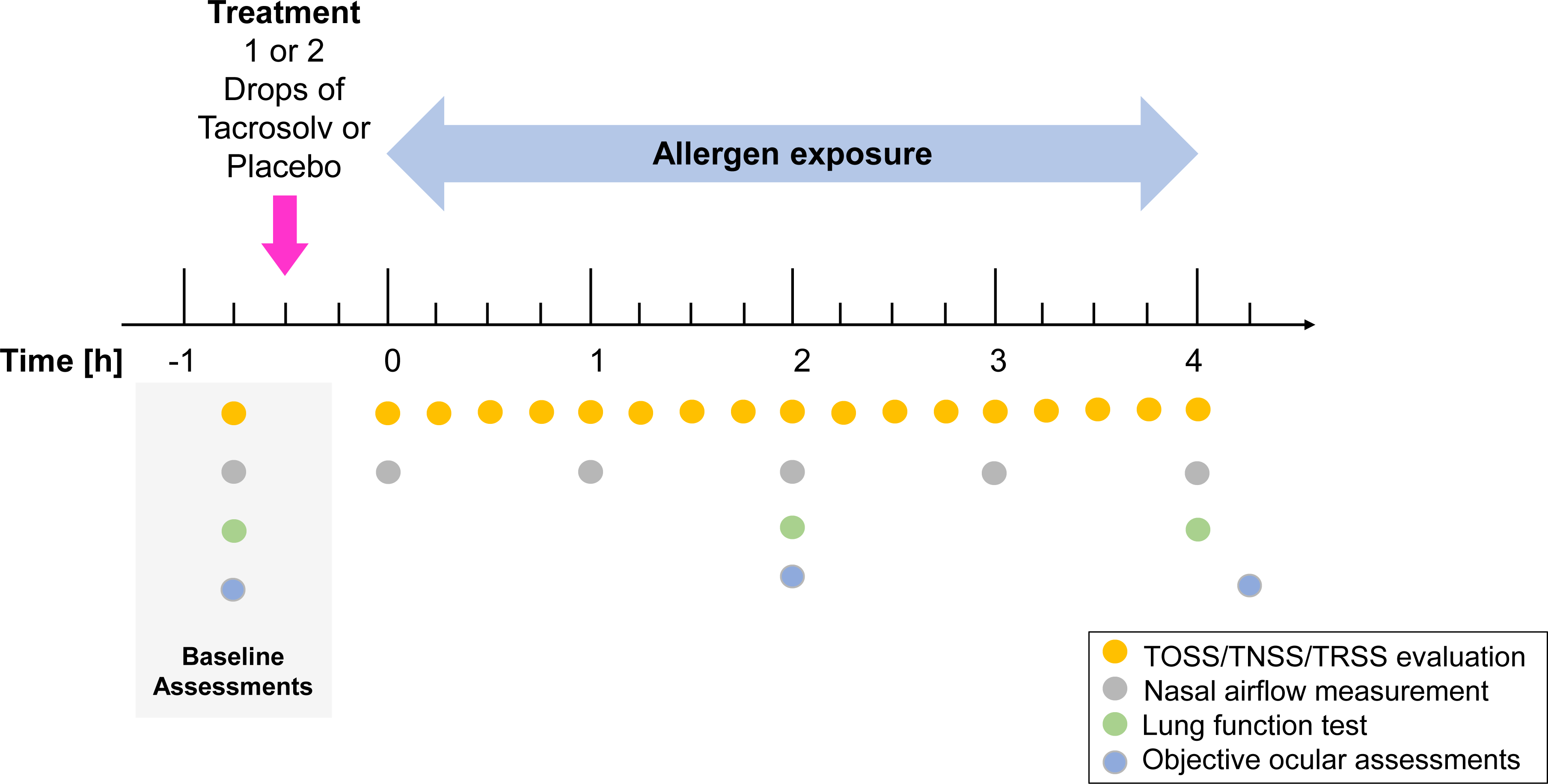
Graphical Abstract of the Clinical Study Panel. A: Study Overview Panel B: Assessments carried out on Day 1 and Day 8 of each treatment block

At visit 4 (day 8 of TMP1), participants returned the study medication kit to the study site staff for compliance evaluation. Baseline symptom scores were assessed, participants received the last dose of their assigned treatment and 30 min after the final treatment, they entered the challenge chamber for another 4-hour allergen exposure. Again, subjective and objective symptom assessments were carried out as described above. After completion of TMP1, a wash-out period of at least 13 days had to be adhered to, allowing complete dissipation of the previous treatment. Subsequently, participants crossed over to their respective next treatment period. Visits 5 and 6 were conducted in an analogous manner as visits 3 and 4. A follow up visit (visit 7, end of trial visit) was scheduled 1-2 weeks after the final allergen exposure session (visit 6).

Participants were asked to record adverse events (AEs) and use of concomitant medications on the provided form during the entire study.

### Participants

Participants were female and male adults aged between 18 and 65 years of either ethnicity/race, with a documented history of clinically relevant moderate to severe seasonal allergic rhinitis (SAR) with rhinoconjunctivitis for the previous two years. Participants were selected from the VCC database and had to satisfy study inclusion and exclusion criteria in order to be enrolled into the study.

The key inclusion criterion was a moderate to severe response to approximately 1800 grains/m^3^ of standard grass pollen in the VCC, defined as TOSS of at least 4 (out of maximum 12) within the first two hours in the challenge chamber, with at least one single ocular symptom scored ≥2 (“moderate”) at least twice during the first two hours. In addition, participants had to fulfill the following inclusion criteria: a positive Skin Prick Test (SPT) response (wheal diameter at least 3 mm larger than diluent control) to grass pollen SPT solution (standard Allergopharma); positive serum specific IgE against recombinant major allergen components of the used grass pollen e.g., g6 (specific CAP IgE ≥0.70 kU/L); and a forced expiratory volume in 1 second (FEV1) of at least 80% of predicted value (ECCS). Key exclusion criteria were uncontrolled or moderate to severe asthma; pregnancy or lactation; smoking; use of contact lenses; previous successful or ongoing treatment with any allergen-specific immunotherapy; symptoms of or treatment for any clinically relevant chronic, systemic or ocular disease affecting the immune response. Female participants of child-bearing potential were required to use birth control.

### Randomization and Blinding

In total, 64 eligible patients were planned to be enrolled into the study. Randomization numbers were allocated to the study participants in ascending order of their screening numbers at Visit 3 (TMP 1). Treatment allocation was based on a cross-over randomization with balanced blocks. All personnel involved in the study, including investigators, site personnel, and sponsor’s staff were blinded to the medication codes. Un-blinding at study end was done after database lock.

### Interventions and Procedures

Tacrosolv is an aqueous solution of 50 µg/ml tacrolimus monohydrate (0.005%). All other components of Tacrosolv except for tacrolimus are classified as excipients and suitable for both ocular and nasal applications; since they have either already been used as excipients in ophthalmic market products and/or have GRAS (“Generally Recognised As Safe”) status.^9^

Sterile buffered saline solution with propylene glycol was used as placebo.

Participants received their first treatment (high or low dose Tacrosolv or placebo) approximately 30 minutes before start of the allergen provocation session on day 1 of the respective TMP. Participants received study medication for the home treatment phase (days 2 to 7 of both TMP1 and TMP2) and continued treatment at home into each conjunctival sac once a day, in the morning, until day 7. On day 8 of the respective TMP, participants applied Tacrosolv or placebo approximately 30 minutes before start of the allergen provocation session.

At the inclusion visit (Visit 2) and on Day 1 and Day 8 of both treatment periods, participants were exposed to standard grass pollen allergen mixture (1800 grass pollen grains/m^3^) in the VCC for 4 hours using a validated method.^10,11^ The challenge agent was a qualitatively and quantitatively defined mixture of four grass pollen species (Timothy, Orchard, Perennial rye and Sweet vernal grass) (Allergon SB, Sweden). Air temperature (24°C), humidity (40%) and allergen load were constantly monitored and maintained. During the 4 hours exposure, subjective ocular, nasal and respiratory symptoms (TOSS, TNSS and TRSS) were recorded every 15 minutes. TOSS is the sum of “ocular redness”, “ocular itching”, “watery eyes” and “gritty feeling”. TNSS is the sum of the symptoms “nasal congestion”, “rhinorrhea”, “itchy nose” and “sneezing”. TRSS is the sum of the symptoms “cough”, “wheeze”, “dyspnea”. Each individual symptom was scored on a 4-point categorical scale from 0 to 3, with 0= complete absence of symptom, 1=mild, 2=moderate and 3=severe.

Lung function was assessed using a Piston spirometer for forced expiratory volume in one second (FEV1) and forced vital capacity (FVC) before, after 2h and at the end of the 4-hour allergen exposure. Nasal airflow was measured by active anterior rhinomanometry (AAR) at a pressure difference of 150 Pascal across the nasal passages (sum of the right and left nostril values) at baseline (45 min before exposure start) and every 60 minutes during exposure.

Objective ocular assessments carried out before and after the allergen challenge session included tear film break-up time (TBUT) measurement, staining of the conjunctiva with lissamine green and of the cornea with fluorescein to evaluate epithelial and corneal damage, evaluation of conjunctival chemosis, lid-parallel conjunctival folds (LIPCOF), of conjunctival redness, eyelid edema and conjunctival papillae with slit-lamp biomicroscopy, and assessment of intraocular pressure (IOP) with a tonometer.

Female participants of child-bearing potential in addition had a urine pregnancy test done at screening and on Day 1 of each treatment period.

### Endpoints

The primary efficacy endpoint was the mean TOSS on day 8, calculated as the mean of TOSS measured every 15 minutes during the pollen allergen exposure.

The key secondary endpoint was the onset of action of either dose of Tacrosolv during the first allergen exposure, defined as first time point when the TOSS difference between active treatment and Placebo was p<0.05. Additional secondary efficacy endpoints were changes in ocular redness image score assessed by the investigator, TNSS, TRSS, nasal airflow assessed by active anterior rhinometry (AAR).

Safety endpoints were frequency, severity, seriousness, and causality of adverse events (AE), lung function (FEV1), vital signs and findings of ocular examinations at screening (V1) and throughout the study (V2-V7), as well as findings of physical examinations, laboratory blood analysis, ECG at Screening (V1) and at the Follow Up Visit (V7).

Objective ophthalmic assessments (eye lid edema, non-invasive first tear film break-up time (NIF-BUT), chemosis, conjunctival papillae, LIPCOF) served as readouts for both efficacy (comparison placebo vs. Tacrosolv treatment) and safety (comparison screening vs. follow-up).

### Sample Size Calculation

Sample size calculation was based on the minimum clinically relevant TOSS difference, which was estimated at about 1 point based on a previous study on solubilized budesonide.^12^ Expecting a mean difference of 1.2 points with a standard deviation of 2.2 (untreated = 8, test = 6.8, effect size d=0.55 and a power = 80%) for each dose group, a total of n=54 participants were needed at an alpha level p=0.05. Considering the dropout rate of 10-15% and 30-40% screening failures, up to 107 participants needed to be screened to randomize about 64 subjects and to obtain evaluable data from at least 54 participants at the end of the trial.

### Statistical Analysis

The final analysis including unblinding was performed on data having been documented as meeting the cleaning and approval requirements defined in the SAP, and after the finalization and approval of the SAP document.

The following 3 analysis populations were defined for this study:

i. Full Analysis Set (FAS), which comprises all participants to whom study drug has been assigned by randomization, analyzed following the intent-to-treat (ITT) principle, i.e., according to the treatment that has been assigned at randomization.
ii. Per-protocol set (PPS), which comprises all participants in the FAS who did not have any clinically important protocol deviations.
iii. Safety set, which comprises all participants who received the investigational product or placebo; used for all safety analyses including vital signs, laboratory data and AEs.

All attempts were made to collect all data as per protocol. Missing or invalid data were not replaced nor extrapolated. Outliers were not excluded from the primary analysis.

For the primary efficacy analysis, a 95% confidence interval was calculated for the mean difference between the active treatment and placebo from a two-sided paired t-test. Superiority of Tacrosolv versus Placebo was to be assumed if the upper limit of the confidence interval did not exceed 0. The FAS was the primary analysis population for the primary efficacy variable.

Secondary efficacy variables were analyzed in an explorative sense. Statistical tests and corresponding p-values were regarded as descriptive and not as tests of hypotheses.

The analysis of baseline and demographic characteristics was subject to descriptive analyses. Safety endpoints were analyzed in the safety set. Adverse events were summarized descriptively. Phase-effects were tested using Wilcoxon tests for both placebo and Tacrosolv. Carry-over effects were tested using ANOVA. The normal distribution was checked using the Shapiro test. If normal distribution was assumed, the paired t-test was used for the group comparison, otherwise the paired Wilcoxon test was used. Confidence intervals are based on t-distributions. Significance level was set to alpha=5%. R version 4.0.3 was used for all statistical analyses.

## Results

### Patient Disposition and Baseline Characteristics

**Figure 1** outlines the study design (**Panel A**) and the assessment carried out on Day 1 and Day 8 (**Panel B**). The study was conducted between December 2020 and April 2021. A total of 93 participants with grass pollen allergy were screened after giving informed consent. Of these, 64 participants complied with all inclusion and exclusion criteria and were randomized to one of four treatment groups, thus constituting the Safety set and the FAS. One participant in the high dose group was lost to follow-up after Day 1, one participant in the low dose group withdrew from the study due to an adverse event not related to the study treatment, and one participant was classified as non-responder after not developing any significant ocular symptoms during the first two hours of the allergy exposure on day 1. Hence, 61 subjects completed the study as per protocol and are comprised in the PPS (**Figure 2**).

**Figure 2.**
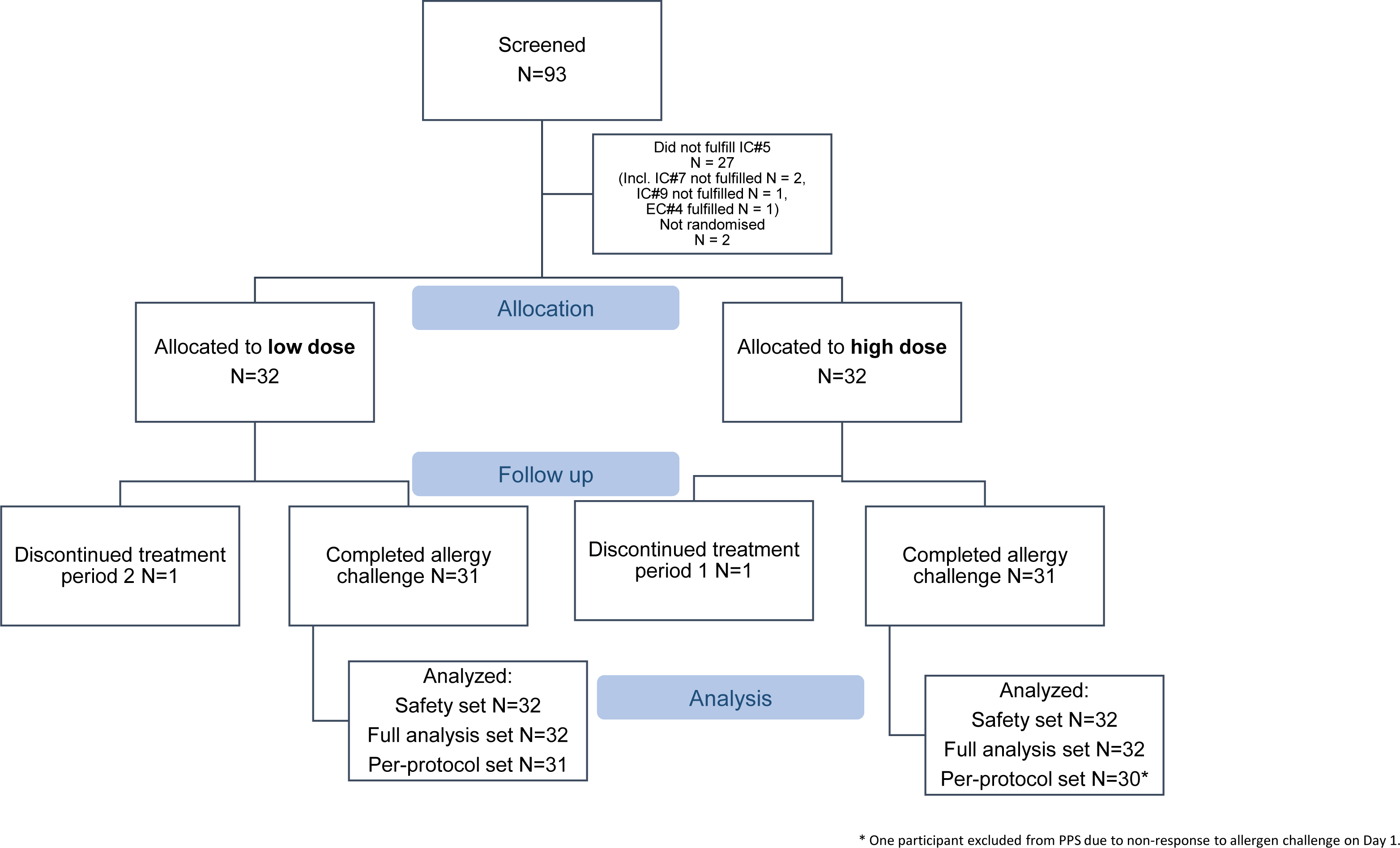
– Version 2: CONSORT Flow Chart.

Demographic characteristics are summarized in **Table 1**. 59% of the participants were females and 41% were males. Participants were aged between 19 and 57 years, with a mean of 32.4 years.

**Table 1:** Demographic Characteristics (Safety set)

|  | Statistics | All participants (N=64) |
| --- | --- | --- |
| Sex |  | 644 |
| Female | n (%) | 38 (59.4 %) 645 |
| Male | n (%) | 26 (40.6 %) 646 |
| Age [Years] | Mean | 32.42 647 |
|  | SD | 9.54 648 |
|  | Min/Max | 19.00/57.00 649 |
| Ethnicity |  | 650 |
| Caucasian | n (%) | 58 (90.6 %) 651 |
| Asian | n (%) | 1 (1.6 %) 652 |
| Hispanic | n (%) | 1 (1.6 %) 653 |
| Other | n (%) | 4 (6.3 %) 654 |
| BMI [kg/m<sup>2</sup>] | Mean | 23.57 655 |
|  | SD | 3.04 656 |
|  | Min/Max | 17.99/30.61 657 |
| Prior duration of allergic rhinitis [Years] | Mean | 20.5 658 |
|  | SD | 9.5 659 |
|  | Min/Max | 3/43 660 |

n, number; SD, standard deviation.

The mean BMI was 23.6 kg/m^2^. All participants had a documented history of moderate to severe SAR with rhinoconjunctivitis to grass pollen with a prior duration of between 3 and 43 years, on average 20.5 years.

### Efficacy

All efficacy results are shown for the FAS. Results for the PPS were similar as for the FAS.

The primary efficacy endpoint was the mean TOSS, calculated as the mean of all TOSS assessments carried out at 15 minutes intervals during the 4-hour grass pollen allergen exposure on day 8. As shown in **Figure 3, upper panel**, there was no statistically significant difference in mean TOSS between the active treatment group versus the placebo group for either high dose or low dose of Tacrosolv on Day 8. With a mean difference of Placebo - Tacrosolv of 0.31, 95% CI [−0.32;0.94], p = 0.328 (paired t-test) in the high dose group and of −0.24, 95% CI [−1.04;0.56], p = 0.54 (paired t-test) in the low dose group, superiority of Tacrosolv over placebo in terms of TOSS on Day 8 could not be stated for either dose group.

**Figure 3.**
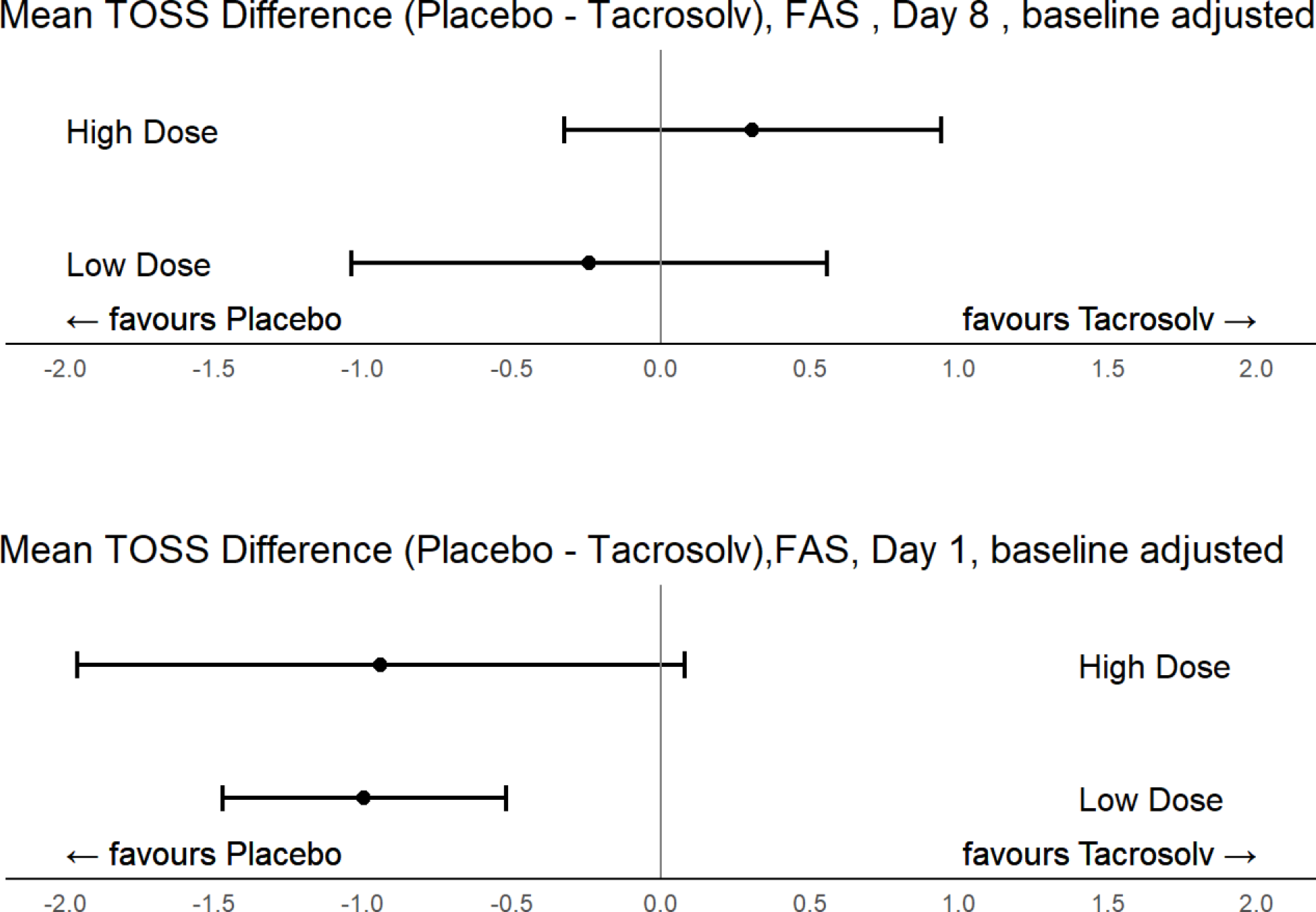
Primary endpoint analysis: Mean and 95% CI for difference of treatments over the entire allergen exposure duration (0-4h), for the FAS on Day 8 (upper panel) and Day 1 (lower panel) Day 8: High dose: mean difference of Placebo -Tacrosolv=0.31, 95% CI [−0.32;0.94], p = 0.328 (paired t-test). Low dose: mean difference of Placebo -Tacrosolv= −0.24, 95% CI [−1.04;0.56], p = 0.54 (paired t-test). Day 1: High dose: Mean difference of Placebo -Tacrosolv= −0.94, 95% CI [−1.96;0.08], p = 0.069 (paired t-test). Low Dose: Mean difference of Placebo -Tacrosolv= -1.00, 95% CI [−1.47;-0.52], p < 0.001 (paired t-test).

On Day 1 (**lower panel**), the mean TOSS difference between active treatment and placebo was similar for the low dose and the high dose group. However, in the high dose group, the mean TOSS difference between Tacrosolv and placebo rose by 1.25 symptom points from −0.94 on Day 1 and to 0.31 on Day 8. Even though the difference was not statistically significant on either day, it showed a trend towards improvement over time.

Since the low dose of Tacrosolv failed to show any beneficial effect in any of the measured parameters, we will in the following focus on the results for high dose Tacrosolv treatment.

The key secondary endpoint was the onset of action of Tacrosolv after the first treatment during the first allergen exposure on Day 1). On Day 1, the mean TOSS was higher in the high dose Tacrosolv group than in the placebo group at all timepoints, reaching a peak value of 6 at timepoint 3h30min and plateauing around 6 for the remaining time (**Figure 4, left panel**). However, on Day 8, the mean TOSS in the high dose Tacrosolv group reached a plateau of only 4 already at timepoint 1h45 min and showed no further increase and only a small range of fluctuation for the remaining duration of the allergen exposure, with the between-groups difference of mean TOSS becoming significant with p<0.05 at timepoints 3:45h and 4:00h (**Figure 4, right panel**). The mean differences at these timepoints exceeded the minimum clinically relevant TOSS difference that was defined as 1 point before study start. The time course of TOSS in the placebo group was the same on Day 1 and Day 8, indicating a high reproducibility of subjective ocular symptoms and no effect of the 8 days placebo treatment.

**Figure 4:**
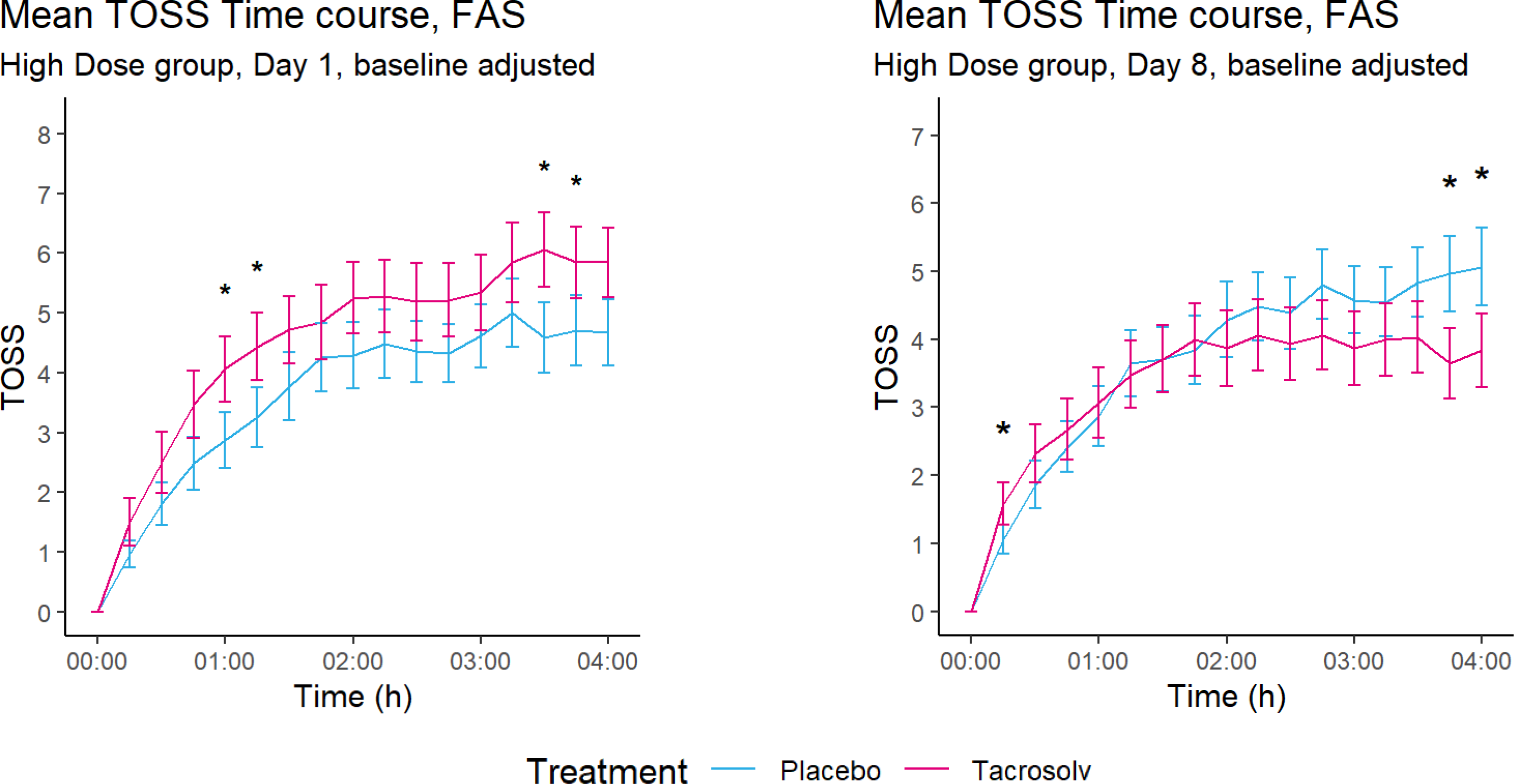
Time course of baseline-adjusted mean Total Ocular Symptom Score (TOSS), full analysis set, high dose group, Day 1 (left panel) and Day 8 (right panel) Day 1: N=31 for placebo, N=32 for Tacrosolv; Day 8: N=31 for both groups. Error bars indicate SEM. * p ≤ 0.05.

When expressing Day 8 mean TOSS as percentage of the Day 1 mean TOSS, it became obvious that only high dose Tacrosolv treatment led to a significantly reduced TOSS on Day 8, compared to Day 1 (**Figure 5, right part of bar chart**). In contrast, there was no difference in mean TOSS between Day 1 and Day 8 in the placebo group (**Figure 5, left part of bar chart**).

**Figure 5:**
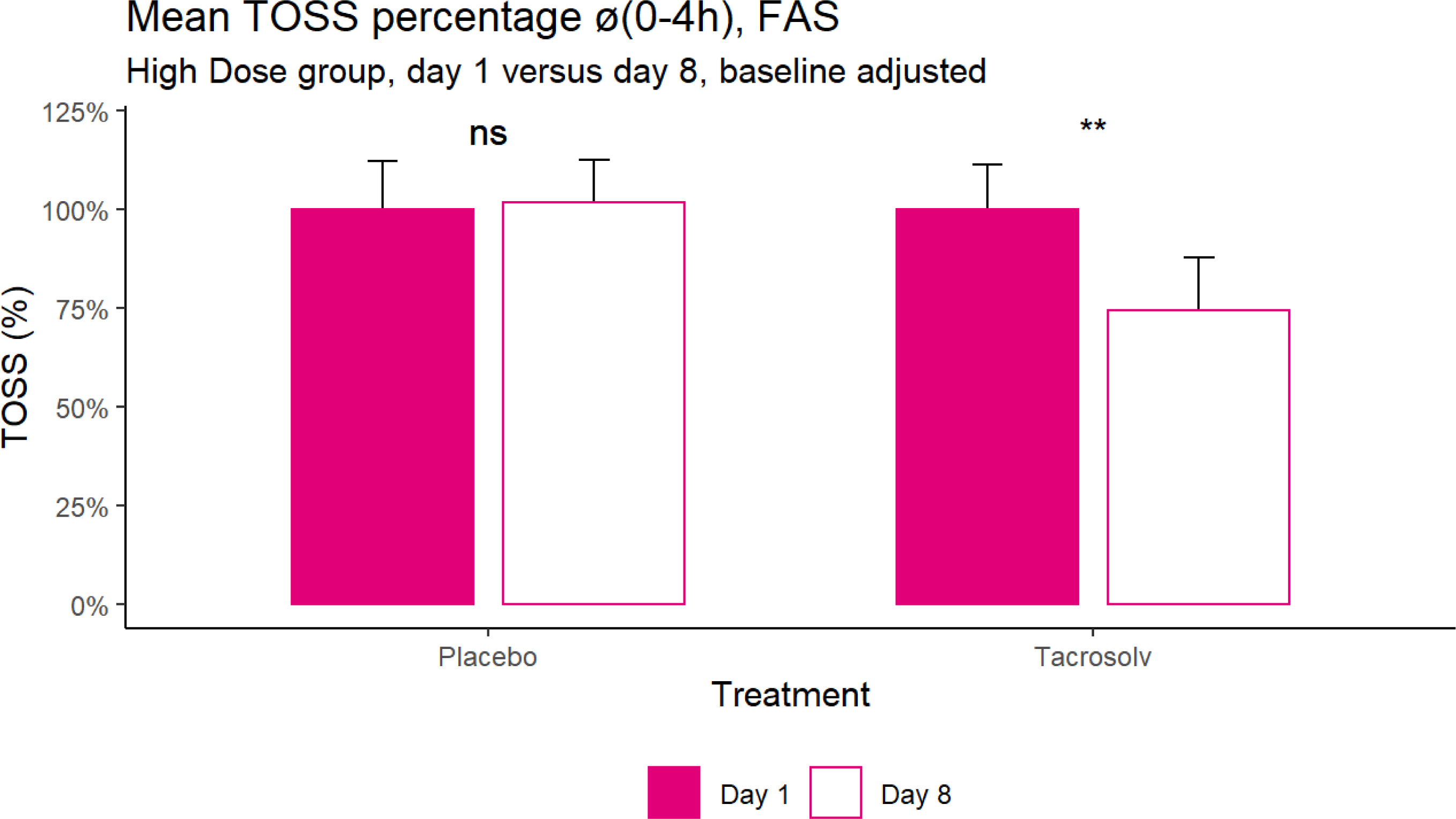
Mean percentage of Total Ocular Symptom Score between 0-4h on Day 1 (100%), compared to Day 8 for the FAS. high dose group. Error bars indicate SEM. ** p ≤ 0.01, assessed by one sample Wilcoxon-test (placebo) and one sample t-test (Tacrosolv).

Time courses of individual TOSS symptoms (itching, redness, watery eyes, gritty feeling) on Day 8 were analyzed post-hoc. As shown in **Figure 6**, treatment with Tacrosolv impacted the three main ocular symptoms associated with allergic conjunctivitis: itchy eyes, redness, and watery eyes. The difference in redness and watery eyes in favor of Tacrosolv became statistically significant towards the end of the allergen exposure. ‘Gritty feeling’ did not contribute to the effect of Tacrosolv on cumulative TOSS.

**Figure 6:**
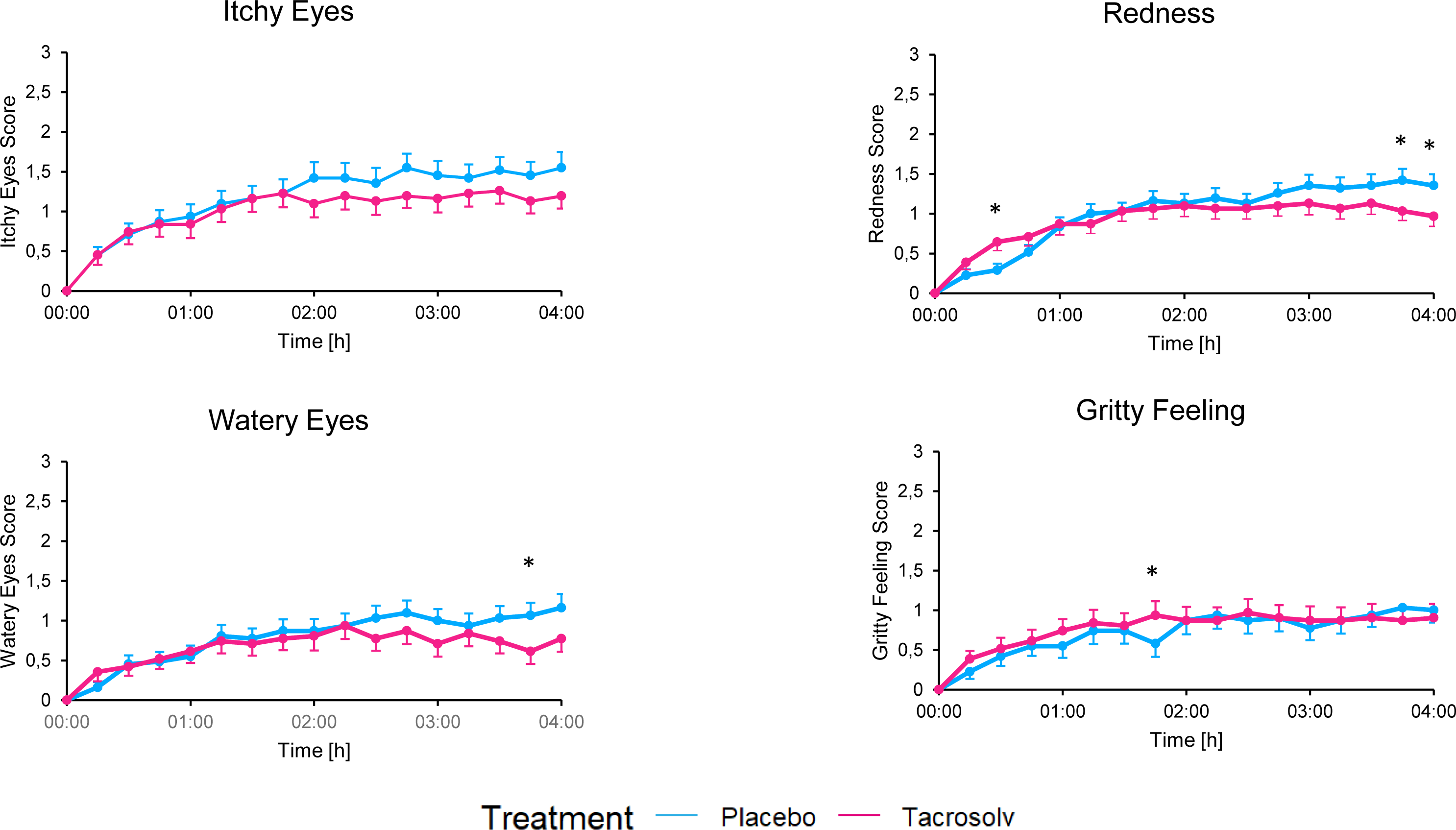
Time course of mean individual, baseline adjusted TOSS symptoms on Day 8, full analysis set, high dose group. N=31 for both groups. Error bars indicate SEM. * p ≤ 0.05.

Interestingly, Tacrosolv treatment benefitted not only ocular, but also nasal symptoms of allergic rhinoconjunctivitis. **Figure 7** shows differences in mean TNSS between high dose Tacrosolv and placebo on Day 1 (**Panel A**) and Day 8 (**Panel B**). On both days, the difference was in favor of Tacrosolv, with a p value for the difference of 0.061 on Day 1 and of 0.034 on Day 8. Time courses of mean TNSS over the 4-hour allergen exposure on Day 1 (**panel C**) and Day 8 (**panel D**) show that the difference in mean TNSS between high dose Tacrosolv and placebo became statistically significant at later timepoints, with 2h30min and 1h45min being the earliest timepoints with significant TNSS difference between treatment groups on Day 1 and Day 8, respectively.

**Figure 7:**
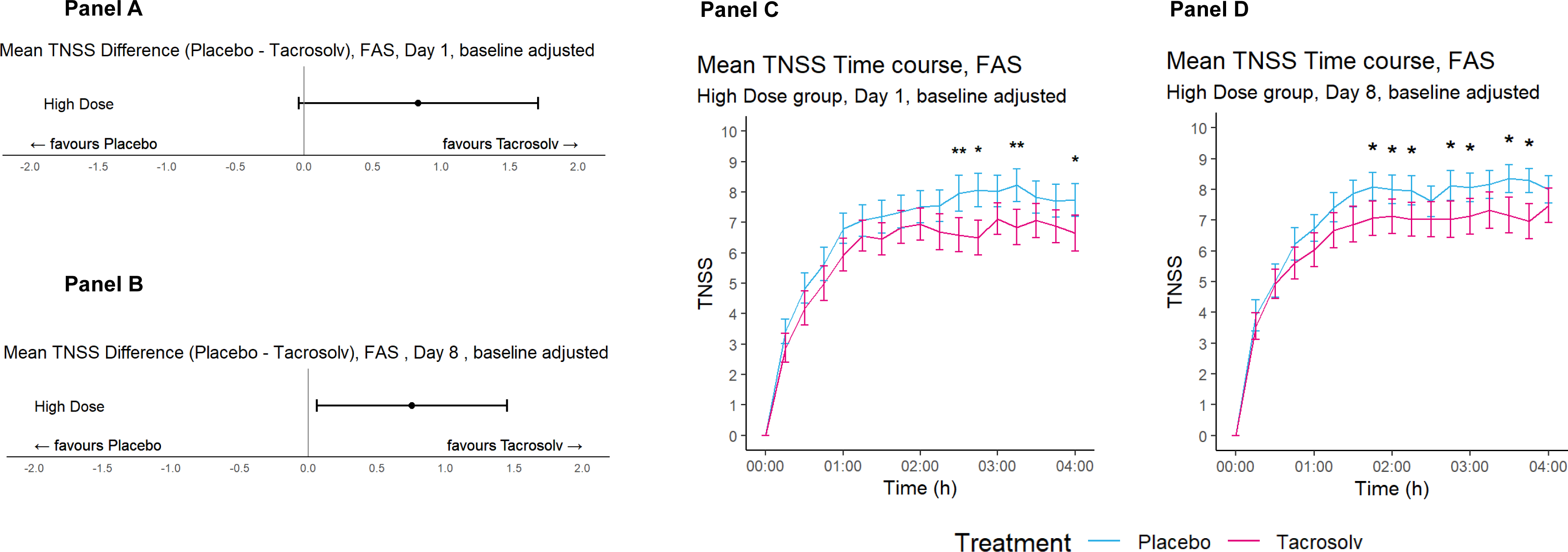
Total Nasal Symptom Score (TNSS) on Day 1 and Day 8 for the full analysis set. Day 1: N=31 for placebo, N=32 for Tacrosolv; Day 8: N=31 for both groups. **Panel A+B: Mean difference between treatments for the FAS on Day 1 (panel A) and Day 8 (Panel B).** Day 1: High dose - mean difference of Placebo - Tacrosolv = 0.83, 95% CI [−0.04;1.71], p = 0.061 (paired t-test). Day 8: High dose - mean difference of Placebo - Tacrosolv = 0.76, 95% CI [0.06;1.45], p = 0.034 (paired t-test). **Panel C+D: Time course of the TNSS, high dose group, Day 1 (Panel C) and Day 8 (Panel D).** Error bars indicate SEM. * p ≤ 0.05, ** p ≤ 0.01

No marked differences between Tacrosolv and placebo groups were observed for ocular redness image score, TRSS and AAR on Day 1 and Day 8. Objective ophthalmic assessments did not reveal any clinically significant findings (data not shown).

No phase-effect was found for Placebo (p-value > 0.05) and Tacrosolv (p-value > 0.05). No carry-over effect was observed (p-value > 0.05).

### Safety and Tolerability

AEs reported throughout the study are summarized for the safety set in **Table 2**. No serious AE, no life-threatening (grade IV) AE and no death occurred throughout the study. A total of 57/64 (89%) participants reported any AE, of which 29 were in the high dose group and 28 in the low dose group. One patient withdrew from the study prematurely due to a non-treatment related adverse event.

20/64 (31%) participants reported at least one AE during the placebo treatment, 55/64 (86%) participants during the active treatment phase with Tacrosolv and 18/64 (28%) participants in both study phases. Severe (grade III) AE occurred in 12/64 (19%) participants during the study (7 were in the high dose group and 5 in the low dose group), and 6 participants required medication in form of artificial tears for treatment of their severe AE(s). 11/64 (17%) participants reported severe AEs during the active treatment phase, and 1 during the placebo phase.

**Table 2:** Overview over adverse events recorded during the study (Safety Set)

| Participants with | Placebo |  | Tacrosolv |  | Placebo and Tacrosolv |  | Total number of patients with AE |  |
| --- | --- | --- | --- | --- | --- | --- | --- | --- |
|  | N | % | N | % | N | % | N | % |
| At least one AE | 20 | 31.25 | 55 | 85.94 | 18 | 28.12 | 57 | 89.06 |
| At least one AE, mild (grade 1) | 16 | 25.00 | 32 | 50.00 | 7 | 10.94 | 41 | 64.06 |
| At least one AE, moderate (grade 2) | 4 | 6.25 | 24 | 37.50 | 3 | 4.69 | 25 | 39.06 |
| At least one AE, severe (grade 3) | 1 | 1.56 | 11 | 17.19 | 0 | 0.00 | 12 | 18.75 |
| At least one serious AE, any | 0 | 0.00 | 0 | 0.00 | 0 | 0.00 | 0 | 0.00 |
| At least one AE leading to early termination | 0 | 0.00 | 1 | 1.56 | 0 | 0.00 | 1 | 1.56 |

Note: Participants are counted once for each category regardless of the number of events. Percentages are calculated in relation to total number of participants in the Safety set (N=64).
AE, adverse event.

A total of 174 AEs were reported during the entire study, of which 30/174 (17%) AEs occurred during the placebo phase and 144/174 (83%) during the active treatment phase. 158 out of 174 (91%) AEs were eye disorders. **Supplemental Table S1** shows AEs by system organ class (SOC) and preferred term (PT). 141 AEs overall were classified by the investigator as probably or possibly treatment-related, the majority of which (129/141, 91%) occurred during the active treatment phase. All adverse events finally were resolved by study end.

Lung function measurements, blood pressure, electrocardiograms and laboratory blood analyses all showed a stable course during the study. No substantial deviations or clinically significant abnormalities were reported. No clinically relevant changes from baseline or relevant differences between treatment groups were observed for any of the analyzed safety parameters.

For ophthalmic parameters (staining of the conjunctiva and cornea, slit lamp microscopy (eye lid edema), fundoscopy, intraocular pressure, chemosis nasal and temporal, conjunctival papillae, LIPCOF temporal, NIF BUT), similar results were observed in the treatment and placebo groups and minor abnormalities were resolved at follow up.

## Discussion

In this proof-of-concept phase II study, we have demonstrated that ophthalmic administration of Tacrosolv, an aqueous solution of 0.005% tacrolimus, applied at a dose of 5 µg/eye/day (“high dose”) over a course of 8 days significantly alleviates ocular and nasal symptoms of pollen allergy in adults with a history of allergic rhinoconjunctivitis.

The high dose Tacrosolv failed to reduce TOSS on Day 1, presumably based on a short-term adverse reaction of a stinging or burning sensation that is well known for tacrolimus^13–15^ and that obscures the beneficial, immune suppressive effect at the start of treatment. Such an instillation site discomfort is also frequently reported for other immunomodulatory ocular medications like cyclosporine or lifitegrast.^16^

In contrast to ocular symptoms, nasal allergy symptoms (like nasal congestion, rhinorrhea, itching and sneezing) were alleviated immediately after the first dose of Tacrosolv. This substantiates the hypothesis that the observed lack of efficacy of Tacrosolv on ocular symptoms on Day 1 may most probably be due to initial, transient adverse effect of tacrolimus that was limited to the site of administration. This transient adverse reaction which is known and commonly reported for topical ophthalmic tacrolimus application.^13–15,17^

It is known that nasal allergen exposure can lead to ocular symptoms,^18,19^ and that treatment of nasal allergy symptoms, e.g. using intranasal corticosteroids, can diminish ocular symptoms.^20–22^ We have previously demonstrated this effect for our Budesolv nasal spray, an aqueous formulation containing solubilized budesonide.^12^

This nasal-ocular relationship is thought to be mediated via a direct and an indirect route.^23^ The direct relation occurs via the nasolacrimal duct that connects the lacrimal sac of the eye with the inferior meatus of the nasal cavity. Along this duct, allergens and allergy mediators drain along with tears into the nasal cavity. As the flow of secretion is from the eye to the nose, it is plausible that ocular drugs travel through the nasolacrimal duct and take effect in the nasal cavity. In addition, an indirect mutual connection between nasal and ocular allergic symptoms may occur through a lymphatic and/or neurogenic pathway.^24,25^ Although the nasal-ocular relationship has mostly been studied in the directionality from nose to eye, there is also evidence for anti-allergic treatment of the eye having an effect in the nose.^26^ Our study confirms the bidirectionality of the nasal-ocular relationship.

Lack of any differences in ophthalmic parameters (staining of the conjunctiva and cornea, slit lamp microscopy (eye lid edema), fundoscopy, intraocular pressure, chemosis nasal and temporal, conjunctival papillae, LIPCOF temporal, NIF BUT) between screening and follow-up showed that both treatment with low and high dose of Tacrosolv as well as exposure to the allergen in the challenge chamber are safe and do not induce any lasting damages as assessed by the applied methods.

In sum, our data indicate an early treatment onset on nasal allergy symptoms and an attenuation of ocular allergy symptoms after 1 week of treatment. The beneficial effect of Tacrosolv became more pronounced, the longer the duration of allergen exposure lasted. We speculate that the effect may further increase with an extended treatment period of more than 1 week and/or when extending the observation period to more than 4 hours. In addition, it may be more meaningful to assess efficacy after reaching the symptoms plateau, which occurs approximately 2h after start of exposure.

Others have defined the meaningful within-patient change and the between-group meaningful difference for patient-reported ocular itching and redness to be approximately 0.5.^27^ It should be noted that in that study, ocular itching, redness and tearing were scored on a 0-4 scale, in contrast to the 0-3 scale used in our study. Others have defined a threshold of 0.23 units as minimum clinically important difference for the TNSS, i.e., the scoring system we have applied in our study.^28^ With a mean TNSS difference between Tacrosolv and placebo of 0.57 (p = 0.145) units on Day 1 and of 0.69 units (p=0.076) on Day 8, we see a clear trend towards a clinically important difference. Hence, our approach defining 1 point as relevant difference can be considered ambitious and we conclude that the difference in TOSS and TNSS observed towards the end of exposure can be considered clinically meaningful.

The observed effect of Tacrosolv is remarkable, since the total dose of tacrolimus applied in this study was only 10 µg per day (i.e., 5µg per eye per day) for the high dose. Talymus^®^, a tacrolimus ocular suspension (Senju Pharmaceutical Co., Ltd., Osaka, Japan) marketed in Japan and South Korea for treatment of vernal conjunctivitis, contains tacrolimus at a concentration of 0.1%. With a recommended dosing of 2 drops per eye per day, assuming a drop size of 50 µl, Talymus^®^ is used at a maximum daily dose of 100 µg per eye per day, that corresponds to 20-fold the high dose applied in our study. This means that with only 5% of the dose used in Talymus^®^, we have achieved a significant, beneficial effect by the end of the 8 days treatment period. Talymus^®^ is formulated as a suspension of finely dispersed tacrolimus particles. In case of molecules with a low solubility and high permeability, like tacrolimus, the bioavailability is greatly influenced by the rate of particle dissolution and the concentration of molecules in solution while in contact with the ocular tissue. In contrast, Tacrosolv enables the solubilization of tacrolimus and therefore, enhances bioavailability and strongly reduces the administered effective dose of tacrolimus.

The safety and efficacy of tacrolimus has previously been proven for the treatment of severe vernal keratoconjunctivitis in children, who were treated with 0.1% tacrolimus twice daily for up to 18 months.^29^ Moreover, a long term study following patients with severe atopic keratoconjunctivitis (AKC) and vernal keratoconjunctivitis (VKC) who were treated with Talymus^®^ (0.1% tacrolimus suspension) for up to 10 years demonstrated safety and efficacy in long term users.^30^

In general, all clinical studies with tacrolimus in allergic eye disease were using either a suspension or an ointment (reviewed in ^31^). Hence, to the best of our knowledge, we we present the first clinical trial using solubilized tacrolimus.

In Europe and the USA, where Talymus^®^ is not available, immunomodulating therapy of severe inflammatory ocular disease is limited to cyclosporine eye drops, while tacrolimus can currently only be used off-label for ophthalmic indiciations.^32^ Comparisons between cyclosporine and tacrolimus in terms of safety and efficacy have mainly been made in the context of immune suppression after organ and tissue transplantation, where tacrolimus has been found to offer similar efficacy as cyclosporine at 20-50fold reduced concentrations.^33^ Studies comparing cyclosporine and tacrolimus eye ointment for the treatment of refractory vernal keratoconjunctivitis have found that tacrolimus demonstrated similar or superior efficacy in reduction of inflammatory symptoms as well as patient compliance,^34,35^ and propose tacrolimus as safe alternative for treatment of cyclosporine-refractory vernal keratoconjunctivitis.^36^

An additional benefit of tacrolimus is that it helps reducing corticosteroid use,^37^ which up to this day is commonly used to treat inflammatory eye disease despite their potential for severe side effects. Tacrolimus treatment efficacy in allergic ocular diseases enabled complete weaning in 50% of patients previously using topical steroids.^30,38^

Severe systemic adverse reactions like hyperglycemia, nephrotoxicity, neurotoxicity, weight loss, liver damage, and diarrhea, which have been reported after systemic use of tacrolimus after bone marrow transplantations,^39,40^ are unlikely to occur upon topical use of Tacrosolv and other tacrolimus-containing eye drops, because the percentage of tacrolimus reaching the bloodstream with twice daily topical use is very low.^41,42^

In contrast to allergic rhinoconjunctivitis which is caused by activation of mast cells, the most severe forms of allergic ocular disease, such as vernal and atopic keratoconjunctivits, involve predominantly T lymphocytes.^43^ Tacrolimus acts on T-cells by disrupting calcium-dependent signaling events and subsequently inhibiting T-cell activation, differentiation and cytokine production,^43,44^ and it is thought that tacrolimus inhibits T-cells even more effectively than mast cells (^3,45–47^ and our own unpublished data). Therefore, Tacrosolv may be even more effective in T-cell mediated ocular diseases than in allergic rhinoconjunctivitis.

One major upside of our study is the setting for eliciting allergy symptoms. Many studies on anti-allergic eye drops apply the conjunctival allergen challenge (CAC), also called conjunctival allergen provocation test (CAPT) or conjunctival provocation test (CPT).^48,49^ In that approach, the allergen is applied directly to the conjunctival mucosa to trigger an allergic response. It is used in clinical practice to determine which allergen trigger specific symptoms, and in clinical research to investigate treatments. In contrast to the CAC, where the initially applied allergen concentration in the eye is quickly diluted and cleared from the mucosa by lacrimation and blinking, use of an environmental challenge chamber enables continuous exposure to the air-dispersed allergen over several hours. In fact, several validation studies demonstrated the reproducibility and specificity of symptoms induced by ECCs,^50,51^ and showed a good correlation between ocular symptoms elicited by ECC and those assessed during natural exposure.^52–54^ Hence, use of the challenge chamber comes very close to allergen exposure in the real world, but with the added benefit of consistent exposure conditions across participants.

In sum, our data demonstrate a beneficial effect on nasal and ocular symptoms of allergic conjunctivitis after 8 days of daily treatment with Tacrosolv 0.005% tacrolimus solution. Hence, our results confirm the therapeutic capacity of tacrolimus for the treatment of allergic eye diseases^43^, and highlight the potential of Tacrosolv as safe and effective treatment option for allergic or inflammatory eye diseases.

## Conclusion

Anti-inflammatory activity of solubilized tacrolimus was observed in subjects suffering from rhinoconjunctivitis at doses as low as 5µg tacrolimus per eye per day.

No major safety concerns were raised during the study. Adverse events were comparable to marketed products containing calcineurin-inhibitors.

## Data Availability

All data produced in the present study are available upon reasonable request to the authors

## Ethics approval and informed consent

The study was conducted in Austria in accordance with the Declaration of Helsinki on Ethical Principles for Medical Research Involving Human Subjects, the International Council for Harmonisation Guideline on Good Clinical Practice, and all applicable local regulatory requirements and laws. The study was approved by the Ethics Committee of the City of Vienna (protocol code TCS_19_02, EK 19-275-1219). Informed consent was obtained from all study participants.

---

**Supplemental Table S1:** Adverse Events by SOC and PTs.

| SOC | PT | Placebo |  | Tacrosolv |  | Total number of AE |  |
| --- | --- | --- | --- | --- | --- | --- | --- |
|  |  | N | % | N | % | N | % |
| Eye disorders |  | 26 | 14.94 | 132 | 75.86 | 158 | 90.80 |
|  | Eye irritation | 13 | 7.47 | 60 | 34.48 | 73 | 41.95 |
|  | Lacrimation increased | 1 | 0.57 | 14 | 8.05 | 15 | 8.62 |
|  | Dry eye | 1 | 0.57 | 14 | 8.05 | 15 | 8.62 |
|  | Eye pruritus | 5 | 2.87 | 9 | 5.17 | 14 | 8.05 |
|  | Ocular hyperaemia | 3 | 1.72 | 9 | 5.17 | 12 | 6.90 |
|  | Photophobia | 1 | 0.57 | 9 | 5.17 | 10 | 5.75 |
|  | Abnormal sensation in eye | 0 | 0.00 | 6 | 3.45 | 6 | 3.45 |
|  | Eye swelling | 1 | 0.57 | 5 | 2.87 | 6 | 3.45 |
|  | Eyelid irritation | 0 | 0.00 | 2 | 1.15 | 2 | 1.15 |
|  | Asthenopia | 0 | 0.00 | 1 | 0.57 | 1 | 0.57 |
|  | Blepharitis | 0 | 0.00 | 1 | 0.57 | 1 | 0.57 |
|  | Eyelids pruritus | 0 | 0.00 | 1 | 0.57 | 1 | 0.57 |
|  | Foreign body sensation in eyes | 0 | 0.00 | 1 | 0.57 | 1 | 0.57 |
|  | Ocular discomfort | 1 | 0.57 | 0 | 0.00 | 1 | 0.57 |
| Nervous system disorders |  | 1 | 0.57 | 6 | 3.45 | 7 | 4.02 |
|  | Disturbance in attention | 0 | 0.00 | 1 | 0.57 | 1 | 0.57 |
|  | Headache | 1 | 0.57 | 4 | 2.30 | 5 | 2.87 |
|  | Migraine | 0 | 0.00 | 1 | 0.57 | 1 | 0.57 |
| Infections and infestations |  | 2 | 1.15 | 2 | 1.15 | 4 | 2.30 |
|  | Conjunctivitis | 1 | 0.57 | 0 | 0.00 | 1 | 0.57 |
|  | Influenza | 0 | 0.00 | 1 | 0.57 | 1 | 0.57 |
|  | Nasopharyngitis | 1 | 0.57 | 0 | 0.00 | 1 | 0.57 |
|  | Pneumonia | 0 | 0.00 | 1 | 0.57 | 1 | 0.57 |
| Immune system disorders |  | 1 | 0.57 | 2 | 1.15 | 3 | 1.72 |
|  | Hypersensitivity | 1 | 0.57 | 2 | 1.15 | 3 | 1.72 |
| Gastrointestinal disorders |  | 0 | 0.00 | 1 | 0.57 | 1 | 0.57 |
|  | Toothache | 0 | 0.00 | 1 | 0.57 | 1 | 0.57 |
| Injury, poisoning and procedural complications |  | 0 | 0.00 | 1 | 0.57 | 1 | 0.57 |
|  | Barotrauma | 0 | 0.00 | 1 | 0.57 | 1 | 0.57 |
| Total |  | 30 | 17.24 | 144 | 82.76 | 174 | 100.00 |

Note: Percentages are calculated related to total number of adverse events (N=174). SOC, System organ class; PT, Preferred term.

